# Do short-term responses to product manipulation health warnings predict subsequent quitting-related behaviours? Findings from an Australian cohort study

**DOI:** 10.1101/2024.10.30.24316410

**Authors:** Emily Brennan, Claudia Gascoyne, Kimberley Dunstone, James F Thrasher, Janet Hoek, Melanie Wakefield, Sarah Durkin

**Author notes:** Corresponding author: Emily Brennan 200 Victoria Parade, East Melbourne 3002, Victoria, Australia.

## Abstract

**Background:** Pictorial health warnings on tobacco packs can stimulate short-term cognitive and emotional responses that lead to quit attempts. We tested novel product attribute health warnings (PAHWs) that corrected misperceptions about harm created by the use of filter-venting, menthol and roll-your-own tobacco. PAHWs uniquely influenced outcomes assessing knowledge of industry manipulation of cigarettes, industry-centric negative emotional responses, and product-specific smoking dissonance. In this study, we examined if these unique short-term responses predicted subsequent quitting-related behaviours.

**Method:** We analysed follow-up data from a between-subjects online experiment that assessed effects of new PAHWs. Participants were randomised to view PAHWs alone (PAHW condition) or with a complementary video (PAHW+Video), and exposure occurred during a baseline session and then repeatedly each day for 7 days. Short-term PAHW responses were measured at 8-day follow-up (N=712). Quitting-related behaviours were measured at 4-week follow-up (N=301). Covariate-adjusted logistic regression models examined associations between short-term PAHW responses and subsequent quitting-related behaviours.

**Results:** Two of the short-term responses – knowledge of industry manipulation of cigarettes, and product-specific smoking dissonance – significantly and positively predicted all three quitting-related outcomes: smoke-limiting micro-behaviours (e.g., foregoing cigarettes), quit attempts, and 7-day sustained abstinence. Additionally, industry-centric negative emotional responses significantly and positively predicted smoke-limiting micro-behaviours and 7-day sustained abstinence, but not quit attempts.

**Conclusion:** PAHWs featuring corrective information about the tobacco industry’s manipulation of tobacco products elicited short-term responses that predicted subsequent engagement in smoke-limiting micro-behaviours, quit attempts, and sustained quit attempts. PAHWs can complement other health warnings featuring the health risks of smoking and may help motivate people who smoke to quit and stay quit.

## INTRODUCTION

Pictorial tobacco health warnings (HWs) play an essential role in encouraging people who smoke to try to quit.[1, 2] To date, tobacco HWs have primarily aimed to improve knowledge about the health consequences of smoking.[3] These types of warnings have been demonstrably effective at increasing awareness of smoking harms, perceived severity of health risks of smoking, and quitting-related help seeking behaviours such as calling a quit support phone line.[1, 2, 4–6] In addition, short-term responses to HWs predict quit intentions and subsequent quitting-related behaviours. For example, various longitudinal cohort surveys have demonstrated that HWs elicit cognitive and emotional responses (i.e., worry about negative outcomes of smoking and thoughts about the harms of smoking) and forgoing of cigarettes, which in turn predict quit intentions and quit attempts.[5, 7–11] More recent experimental research has further demonstrated that short-term responses tapping into the perceived impact of tobacco HWs (e.g., whether the warning discourages people from wanting to smoke, or makes them feel concerned about the health effects of smoking) are strong predictors of subsequent quit intentions and multiple quitting-related behaviours.[12] These findings are consistent with literature on the predictive validity of perceived effectiveness measures used to assess responses to anti-smoking television advertisements.[13]

In contrast to HWs that focus on smoking’s health harms, HWs could also provide corrective information about specific attributes of tobacco products known to mislead people who smoke about smoking harms. Our team recently developed novel product attribute health warnings (PAHWs) that explain how the tobacco industry manipulates their products to produce appealing sensory cues, thereby masking the true underlying harshness of tobacco smoke and dampening sensations associated with harm.[14] For example, tobacco products that contain menthol are commonly perceived to be less harmful than non-menthol products due to the fresh taste and soothing sensations on the throat;[15] yet in reality, products that contain menthol are associated with increased nicotine dependence, increased risk of progression to heavier smoking, and decreased rates of smoking cessation.[16, 17] Similarly, roll-your-own (RYO) tobacco is also commonly misperceived to be less harmful and more natural than tailor-made (TM) cigarettes (also known as factory-made cigarettes).[18–20] However, RYO tobacco contains many harmful additives to improve the palatability of the harsh raw tobacco, such as humectants to retain moisture.[21, 22] A third common tobacco product attribute that misleads consumers about harms is ventilated filters. These filters contain tiny holes to allow the smoke to be diluted with air during inhalation, thereby creating lighter- and smoother-feeling sensations that delude consumers into believing that the smoke is less harmful.[23] Harm misperceptions related to filter-ventilated products are driven by the tobacco industry’s historical marketing of these products as ‘light’ or ‘low-tar’.[24] Tobacco companies responded to bans on these misleading terms by instead using ‘smooth’ and other similar descriptors and light-coloured packaging (e.g., white, silver, gold) to signify reduced harshness and harm,[25–27] and by using lighter-colour variant names in nations where tobacco packaging was standardised to a single colour.[28]

The PAHWs we developed employ imagery and text to provide corrective information that demonstrates how the industry manipulates products to create these misleading sensations. PAHW development involved an iterative process across six sequential studies (described elsewhere[14]), commencing with exploratory qualitative research and building to a final quantitative message testing study that confirmed each of the final PAHWs improved targeted knowledge[14] and performed well against several standard perceived effectiveness constructs, including believability, clarity, understanding and credibility.[29]

Following message development, our team conducted a four-condition experimental study with over 2,500 Australian adults who smoked to assess the effectiveness of the new PAHWs, and whether a video advertisement complementing the PAHWs could augment effects of exposure to PAHWs alone. Compared to a control condition, standard health harm tobacco HWs (newly developed for the study) and the new PAHWs were comparably effective on several key outcomes, including certain negative emotional responses (i.e., worry, discomfort, embarrassment) and message rumination. Exposure to both the PAHWs and complementary video produced additional responses, including greater online information-seeking and inter-personal discussion about the PAHW messages. Importantly, many of these HW responses predict subsequent quitting-related outcomes.[10, 30–33]

Given the novel message content featured on the PAHWs, three new constructs were also found to be unique responses to the PAHWs, with effects above and beyond those observed for standard tobacco HWs. These measures include i) knowledge of industry manipulation of tobacco products (e.g., knowledge that tobacco companies modify cigarettes to change the way the cigarette smoke feels and tastes), ii) industry-centric negative emotional responses (e.g., feeling angry at or deceived by the tobacco industry), and iii) product-specific smoking dissonance (e.g., feeling put off from continuing to smoke their current tobacco product). However, the predictive validity of these three responses that are unique to PAHWs is unknown, so it is unclear whether these measures provide any meaningful indication of the likelihood of subsequent quitting activity.[34]

Our study therefore aimed to extend these earlier findings by examining how well these new short-term outcome measures (knowledge of industry manipulation, industry-centric negative emotional responses, and product-specific smoking dissonance) predict subsequent quitting-related behaviours, including engagement in smoke-limiting micro-behaviours (e.g., foregoing cigarettes), quit attempts and sustained quit attempts (of at least seven days) in the four weeks following initial exposure.

## METHODS

### Study design

The current study is a secondary analysis of data extracted from a four-arm between-subjects online experimental study of tobacco health warnings, where participants were randomly assigned to one of four conditions: i) no health warnings (control; warnings for over-the-counter pharmaceutical medications), ii) refreshed Standard HWs featuring imagery and text about the health harms of smoking, iii) Product Attribute HWs featuring imagery and text about product attributes known to mislead people who smoke about harms, and iv) PAHWs + a complementary video advertisement (PAHW+Video). Participants completed a baseline survey during which they were exposed to their assigned stimuli. Participants were then able to opt-in to complete up to seven daily repeated exposure tasks (RETs), during which they were potentially re-exposed to the same stimuli (i.e., re-exposed to one HW on each day that they completed a RET). Follow-up surveys were sent at 8-days and again at 4-weeks. The study methodology has been described in detail elsewhere.[14] The study was approved by the Human Research Ethics Committee of Cancer Council Victoria (IER 1706).

### Sample and procedure

An ISO-accredited data collection agency recruited participants from Australian online non-probability panels via email and collected data between 3^rd^ August and 1^st^ November, 2022. Eligible participants were aged between 18 to 69 years, resided in Australia at the time of the survey, and currently smoked any type of tobacco cigarette at least weekly. Recruitment applied soft quotas for gender and age group (non-intersecting, +/- 5% tolerance) and hard quotas based on the type of cigarettes predominantly smoked (54% TM, 25% menthol, 21% RYO). Participants were offered a reward value of AUD $18 to complete the baseline survey, after which they were offered a reward value up to AUD $16.50 to participate in the seven daily one minute RETs and/or the follow-up surveys sent 8-days and 4-weeks after baseline survey completion. Data collection for both follow-up surveys remained open to allow participants to complete the surveys at a convenient time, thereby limiting potential for loss to follow-up. This methodology resulted in some variation in the actual number of days between surveys, as described in the results. For the current analyses, the analytic sample comprised N=301 participants who were assigned to either the PAHW or PAHW+video condition and who chronologically completed all three surveys.

### Interventions

All participants assigned to the intervention conditions were exposed to seven of 11 possible HWs relevant to their condition, displayed on plain standardised Australian packs, with HW imagery and text on the front, side, and back of pack (or under the flap for RYO pouches). Two examples of the PAHWs are provided in the Supporting Information Figure S1, and a copy of all PAHWs is available from the corresponding author upon request. Participants were shown the entirety of the pack (or pouch) across various enlargeable images, which were shown during the baseline survey and each RET for a minimum of six seconds.

PAHW and PAHW+Video messages focussed on harm misperceptions associated with filter-ventilated cigarettes, menthol tobacco products, and roll-your-own tobacco. All PAHW and PAHW+Video condition participants were shown three PAHWs that informed participants that most of the harm from smoked tobacco products was due to the process of combustion, rather than from additives which are included to mask the harshness of smoke. Participants were also shown two PAHWs relevant to the type of tobacco product they predominantly smoked (tailor-made cigarettes; roll-your-own cigarettes; or menthol/crushball cigarettes), and two PAHWs relevant to products they did not predominantly smoke, mirroring real-world exposure to a diverse range of HWs and conveying that switching to an alternative product would not lower the risk of health consequences.

The video shown to participants in the PAHW+Video condition featured similar messages to the PAHWs regarding product attributes that mislead people who smoke about harm. Participants were twice exposed to a 30-second version at the baseline survey and were potentially re-exposed to a 15-second version during each of the daily RETs.

### Measures

#### Responses to health warnings measured at 8-day follow-up

During the 8-day follow-up survey, participants responded to various measures that utilised Likert-type scales ranging from 1 (“strongly disagree”) to 5 (“strongly agree”), which were combined into composite mean scores for analysis. Two measures were combined to measure knowledge of tobacco industry manipulation: i) “tobacco companies process raw tobacco to change the way the cigarette smoke feels and tastes”, and ii) “tobacco companies modify cigarettes to change the way the cigarette smoke feels and tastes” (α=0.807). Industry-centric negative emotional responses comprised agreement that the HWs (plus video for PAHW+Video) made them feel i) angry at tobacco companies, and ii) deceived by tobacco companies (α=0.762).

Five measures were combined to create the product-specific smoking dissonance scale. Three items asked participants to consider how they had felt as they inhaled the smoke from their cigarettes over the past week: i) found it less enjoyable than before, ii) felt more uncomfortable about their smoking than before, and iii) felt more uneasy about their cigarettes than before. Two items asked about the extent to which participants had, in the past week, iv) thought about the harm caused by their tobacco product more than they used to, and v) felt put off from continuing to smoke their current tobacco product (α=0.864).

#### Quitting-related behaviours measured at 4-week follow-up

To measure engagement in smoke-limiting micro-behaviours, participants reported how often over the past month they had i) tried to limit the number of cigarettes they smoked, ii) stubbed or butted out a cigarette before finishing it, and iii) stopped themselves from having a cigarette when they had the urge to smoke. Response options comprised “not at all”, “once or twice”, “several times”, “many times”, and “don’t know / can’t say”. The outcome of interest was having engaged in all three smoke-limiting micro-behaviours at least once in the past month.

Participants were also asked to report how many times, if any, they had tried to quit smoking for at least 24 hours in the past month. Those who reported doing so at least once were classified as having made a quit attempt. Participants who had made a quit attempt were also asked when their last attempt to quit smoking had ended. If participants were still not smoking, they were asked how long it had been since they last smoked, and if participants had resumed smoking, they were asked how long they had stopped smoking for on their last quit attempt. Participants who were still not smoking and had not smoked for at least seven days, as well as participants who were currently smoking but had not smoked for at least seven days during their last attempt to quit, were classified as having abstained from smoking for at least seven days in the past month (7-day sustained abstinence).

#### Demographic and smoking characteristics

At the baseline survey, participants reported their gender, age, highest level of education, whether they held a government-issued health care or pensioner concession card (as an individual-level indicator of socio-economic status), and whether they identified as Aboriginal or Torres Strait Islander. Participants also reported their residential postcode, which identified the socio-economic status of the area they lived in according to the 2021 Index for Relative Socio-economic Disadvantage,[35] as well as regionality (metropolitan or regional).[36]

At the baseline survey, participants reported how many quit attempts they had made in the prior year, the number of TM and RYO cigarettes smoked per day, frequency of e-cigarette use, and whether they were planning to quit smoking in the next 30 days. Participants also reported their frequency of use of different tobacco products (TM, RYO, menthol and menthol crushball cigarettes), and were categorised as predominantly smoking either TM or RYO cigarettes depending on which product they used most frequently, or as a predominant smoker of menthol cigarettes if they currently smoked menthol and/or menthol crushball cigarettes at least daily (regardless of their frequency of use of other tobacco products).

### Statistical analysis

We conducted a series of adjusted logistic regression models to examine associations between each HW response measured at the 8-day follow-up and each outcome measured at the 4-week follow-up. For analysis we combined the sample of participants assigned to the PAHW and PAHW+video conditions (hereafter referred to as the PAHW conditions), given that the predictors of interest were responses to PAHWs and doing so provided a larger analytic sample and therefore greater statistical power. Furthermore, the initial experimental study found that the direction of effects on all outcomes for the PAHW and PAHW+video conditions were the same and there were no statistically significant differences in effect sizes for these two conditions.[14] Participants were excluded from all analyses if they were missing data for any variables used as predictors, outcomes, or covariates.

All reported models included eight covariates in total. Consistent with previous research,[37] baseline intentions to quit in the next 30 days was included as a measure of readiness to quit. The number of days between the 8-day and 4-week surveys was also included based on the likelihood of having engaged in a quitting-related behaviour in the past month increasing as more time elapsed between surveys. Age group (18-39 years; 40-69 years) and baseline number of cigarettes smoked per day were included because these variables are commonly associated with quitting-related behaviours. Predominant product use (TM cigarettes; RYO cigarettes; menthol TM and/or RYO cigarettes) was included because it was associated with loss to follow-up at the 4-week survey. In our study sample, baseline frequency of e-cigarette use (less than monthly; at least monthly) was associated with all three quitting-related outcomes, hence all models included frequency of e-cigarette use as a covariate. In addition, region (metropolitan; regional) was associated with smoke-limiting micro-behaviours and was therefore included in all models pertaining to this outcome, while gender (male; female) was associated with quit attempts and 7-day sustained abstinence and was included in all models pertaining to these two outcomes. Sensitivity analyses also identified that the inclusion of 8-day measures of smoke-limiting micro-behaviours and quit attempts as covariates did not significantly influence the pattern of results, although as expected, effects were slightly attenuated (see Supporting Information; Table S2). This indicates that our main results are not being driven by quitting-related behaviours during the time period between the baseline survey and the 8-day follow-up survey.

For all models, adjusted odds ratios (ORs) are reported. A significance level of *p*<0.05 was used. Data were analysed using Stata MP version 16.0.

## RESULTS

### Participants

1,295 participants completed the baseline survey. 737 (56.9%) of these completed the 8-day follow-up survey, although 25 were excluded due to survey programming error, leaving 712 respondents at the 8-day follow-up. Of those 712 participants, 301 completed the 4-week follow-up survey, representing 23.2% of all participants and 40.8% of those who completed the 8-day follow-up. Among these 301 participants, the mean number of days between completion of the baseline and 8-day follow-up survey was 9.2 days, and the mean number of days between completion of the baseline and 4-week follow-up survey was 33.6 days. Table 1 displays the socio-demographic and smoking characteristics of this sample.

**Table 1.** Socio-demographic and smoking characteristics among the analytic sample of participants who completed the baseline, 8-day follow-up and 4-week follow-up survey.

|  | Analytic sample of participants who completed all three surveys |
| --- | --- |
|  | n (%) |
| *Total* | 301 (100.0) |
| *Age group* |  |
| 18-39 years | 158 (52.5) |
| 40-69 years | 143 (47.5) |
| *Gender<sup>a</sup>* |  |
| Man / male | 135 (44.9) |
| Woman / female | 165 (54.8) |
| Another term | 0 (0.0) |
| Prefer not to say | 1 (0.3) |
| *Highest level of education* |  |
| No tertiary education | 185 (61.5) |
| Tertiary education | 115 (38.2) |
| *Socio-economic area* |  |
| Low (1-40%) | 120 (39.9) |
| Mid (41-80%) | 119 (39.5) |
| High (81-100%) | 62 (20.6) |
| *Geographic region* |  |
| Metropolitan | 212 (70.4) |
| Regional | 89 (29.6) |
| *Aboriginal and/or Torres Strait Islander<sup>b</sup>* |  |
| No | 287 (95.4) |
| Yes | 12 (4.0) |
| Prefer not to say | 2 (0.7) |
| *Health Care Card or Pensioner Concession Card holder* |  |
| No | 200 (66.5) |
| Yes | 101 (33.6) |
| *Predominant product use* |  |
| TM cigarettes | 139 (46.2) |
| RYO cigarettes | 69 (22.9) |
| Menthol cigarettes | 93 (30.9) |
| *Number of cigarettes smoked per day* |  |
| <10 | 147 (48.8) |
| 10+ | 146 (48.5) |
| *Quit attempts in past year* |  |
| None | 136 (45.2) |
| At least once | 147 (48.8) |
| Don't know/can't say | 18 (6.0) |
| *Intention to quit in next 30 days* |  |
| No | 238 (79.1) |
| Yes | 63 (20.9) |
| *Frequency of e-cigarette use* |  |
| Less than monthly | 192 (63.8) |
| At least monthly | 109 (36.2) |
| *Condition* |  |
| PAHW | 158 (52.5) |
| PAHW+Video | 143 (47.5) |

Notes: Proportions are rounded so may not sum to 100.0%. No response was provided for highest level of education for n=1 participants and no response was provided for number of cigarettes smoked per day for n=8 participants.
Abbreviations: TM=tailor-made; RYO=roll-your-own; PAHW=product attribute health warning.

Among participants who completed both the baseline and 8-day follow-up, those who did and did not complete the 4-week follow-up were approximately equivalent in terms of baseline socio-demographic and smoking characteristics (see Supporting Information; Table S1). However, participation in the 4-week follow-up survey differed by predominant product use (TM, RYO or menthol; *p*<0.001), and hence this measure was included as a covariate in all models.

### Quitting-related outcomes

At the 4-week follow-up, 69.4% had engaged in all three smoke-limiting micro-behaviours in the past month, 48.5% had made at least one quit attempt, and 8.3% had made a quit attempt that lasted at least 7 days.

### Predictive validity of responses to the PAHWs

As shown in Table 2, greater knowledge of industry manipulation of cigarettes positively predicted smoke-limiting micro-behaviours (*p*=0.034), quit attempts (*p*=0.013) and 7-day sustained abstinence (*p*=0.011) at the 4-week follow-up survey. Greater industry-centric negative emotional responses positively predicted smoke-limiting micro-behaviours (*p*<0.001) and 7-day sustained abstinence (*p*=0.037), but the association with quit attempts was not significant (*p*=0.090). Greater product-specific smoking dissonance predicted smoke-limiting micro-behaviours (*p*<0.001), quit attempts (*p*<0.001) and 7-day sustained abstinence (*p*=0.003).

**Table 2.** Results from binary logistic regression models assessing predictive validity of PAHW response measures for quitting-related behavioural outcomes.

|  | All three smoke-limiting behaviours<br>(N=283) |  | At least one quit attempt<br>(N=292) |  | 7-day sustained abstinence<br>(N=292) |  |
| --- | --- | --- | --- | --- | --- | --- |
|  | Adj. Odds Ratio<br>(95% CI) | *p*-value | Adj. Odds Ratio<br>(95% CI) | *p*-value | Adj. Odds Ratio<br>(95% CI) | *p*-value |
| Knowledge of industry manipulation of cigarettes | 1.50 (1.03, 2.20) | 0.034 | 1.66 (1.11, 2.47) | 0.013 | 2.85 (1.27, 6.42) | 0.011 |
| Industry-centric negative emotional responses | 1.73 (1.27, 2.36) | <0.001 | 1.30 (0.96, 1.76) | 0.090 | 1.88 (1.04, 3.38) | 0.037 |
| Product-specific smoking dissonance | 2.24 (1.53, 3.27) | <0.001 | 2.66 (1.76, 4.00) | <0.001 | 2.96 (1.44, 6.09) | 0.003 |

Note: All models adjusted for intentions to quit in the next 30 days measured at baseline, number of days between 8-day and 4-week follow-up surveys, age group, number of cigarettes smoked per day, predominant product use, and frequency of e-cigarette use. Models pertaining to smoke-limiting behaviours also adjusted for region, and models pertaining to quit attempts and 7-day point sustained abstinence also adjusted for gender. Participants were excluded from analysis if they were missing data for any variables used as predictors, outcomes, or covariates.

## DISCUSSION

This study examined the predictive validity of short-term responses representing psychosocial pathways that are unique to newly designed PAHWs, including knowledge of industry manipulation of cigarettes, industry-centric negative emotional responses, and product-specific smoking dissonance. All three measures predicted subsequent quitting-related behaviours; these findings support the utility of measuring these short-term responses to assess PAHW effects, while also suggesting that exposure to PAHWs can promote subsequent quitting activity.

Greater knowledge of industry manipulation of cigarettes and product-specific smoking dissonance were statistically significant predictors of all three quitting-related outcomes—smoke-limiting micro-behaviours, quit attempts, and 7-day sustained abstinence. These findings suggest that providing explicit and factual information that increases knowledge of industry manipulation of cigarettes can be a persuasive avenue through which to increase motivation to quit. Previous research has found that the effect of HWs on subsequent quit attempts is mediated through thoughts about the health risks of smoking and worry about negative outcomes of smoking,[8] and hence product-specific smoking dissonance may produce a similar yet more personally-relevant effect, given that the items included in the scale relate specifically to each participant’s preferred product. We also observed that industry-centric negative emotional responses (i.e., feeling angry at or deceived by the tobacco industry) predicted smoke-limiting micro-behaviours and 7-day sustained abstinence, with marginal effects observed for quit attempts. These findings align with previous research, which has identified that anti-industry attitudes and are associated with quit intentions and quit attempts,[38] and tobacco industry denormalization can reduce smoking prevalence and initiation on the population level.[39]

It is possible that while industry-centric negative emotional responses can motivate an individual to take the first steps towards quitting by engaging in smoke-limiting micro-behaviours, these emotional responses may be inherently short-lived and therefore less effective in driving quit attempts, noting that our participants were not re-exposed to the PAHWs at all between completing the 8-day and 4-week follow-up surveys. Previous research has also identified that stronger negative emotional responses to standard HWs predict stronger quit intentions[40] and increases the likelihood of subsequent quit attempts.[10] Therefore, repeated exposure to PAHWs may provide regular reminders and reinforcement of the negative emotions towards the industry, which, if sustained, may promote quit attempts.

Our study findings indicate that knowledge of industry manipulation of cigarettes, industry-centric negative emotional responses, and product-specific smoking dissonance can be reliably used to evaluate the impact of corrective messages featured on PAHWs. These types of corrective messages have been shown to increase quit intentions[41] and can play an important role in motivating people who smoke to quit given that use of certain products that are commonly misperceived to be ‘lower risk’ is associated with lower odds of smoking cessation.[42]

Increasing knowledge about misleading tobacco product attributes may also build public support for policy change to regulate certain product attributes and thereby reduce the appeal and addictiveness of smoking. In addition to HWs, corrective messages can also be delivered via alternative formats such as video-based integrated public communication campaigns. Tobacco control campaigns have increasingly featured corrective messages in recent years, for example as part of ‘The Con That Kills’ campaign developed by Quit Victoria, which aimed to educate people who smoke about how the feeling of inhaling smoke is manipulated,[43] as well as the ‘It’s Not Just’ campaign developed by Tobacco-Free New York State, which aimed to correct misconceptions that menthol is a harmless flavour.[44] The three predictors of quitting behaviours assessed as part of the current study may also be used to evaluate the impact of such communication campaigns. Furthermore, these three predictors also provide discriminant validity given that they were not found to be responses to the standard tobacco HWs in the main experimental study.[14] In line with a recent systematic review of measures used in experimental studies of tobacco HWs, the predictors assessed are multi-item scales, which provides reassurance that these measures are comprehensively portraying the intended constructs.[34]

### Strengths and limitations

Predictors assessed within the current study may be related to one another in more complicated ways than is currently captured, so further mediation analyses of longitudinal data may be able to more precisely elucidate the pathway from HW responses to quitting-related behaviours. Furthermore, all data are self-reported and therefore subject to social desirability bias and recall bias. Since the 4-week follow-up outcomes assessed behaviours in the past month and the predictors were measured at the 8-day follow-up survey, there is a possibility that the outcomes capture behaviours from the one week prior to measurement of the predictors, however sensitivity analysis that controlled for 8-day measures provided reassurance that this aspect of the study design had minimal impact on the findings. The generalisability of study findings is somewhat limited, given that a quota was set to ensure the study sample included an adequate number of adults who smoked menthol cigarettes. While fairly low sample sizes and retention rates were observed (23% of baseline participants and 41% of 8-day follow-up participants completed the 4-week follow-up survey), the study is strengthened by the use of a cohort design allowing for causal inferences. The study is also strengthened by the inclusion of a comprehensive set of covariates, including baseline 30-day quit intentions.

## Conclusion

Taken together, these findings suggest that the unique short-term responses that are elicited by exposure to PAHWs predict subsequent quitting-related behaviours, reinforcing existing evidence suggesting that these types of corrective messages may have a place in a broader suite of health warnings. The unique measures examined as part of the current study – knowledge of industry manipulation of cigarettes, industry-centric negative emotional responses, and product-specific smoking dissonance – can be used to meaningfully assess and evaluate the efficacy of future tobacco control interventions and communication strategies.

## Data Availability

The data underlying this article will be shared upon reasonable request to the corresponding
author.

