## Supplemental Figure 1 (S1) for "Do short-term responses to product manipulation health warnings predict subsequent quitting-related behaviours? Findings from an Australian cohort study"

**Figure S1.** Example Product Attribute Health Warnings (PAHWs). A copy of all PAHWs is available upon request to the corresponding author.
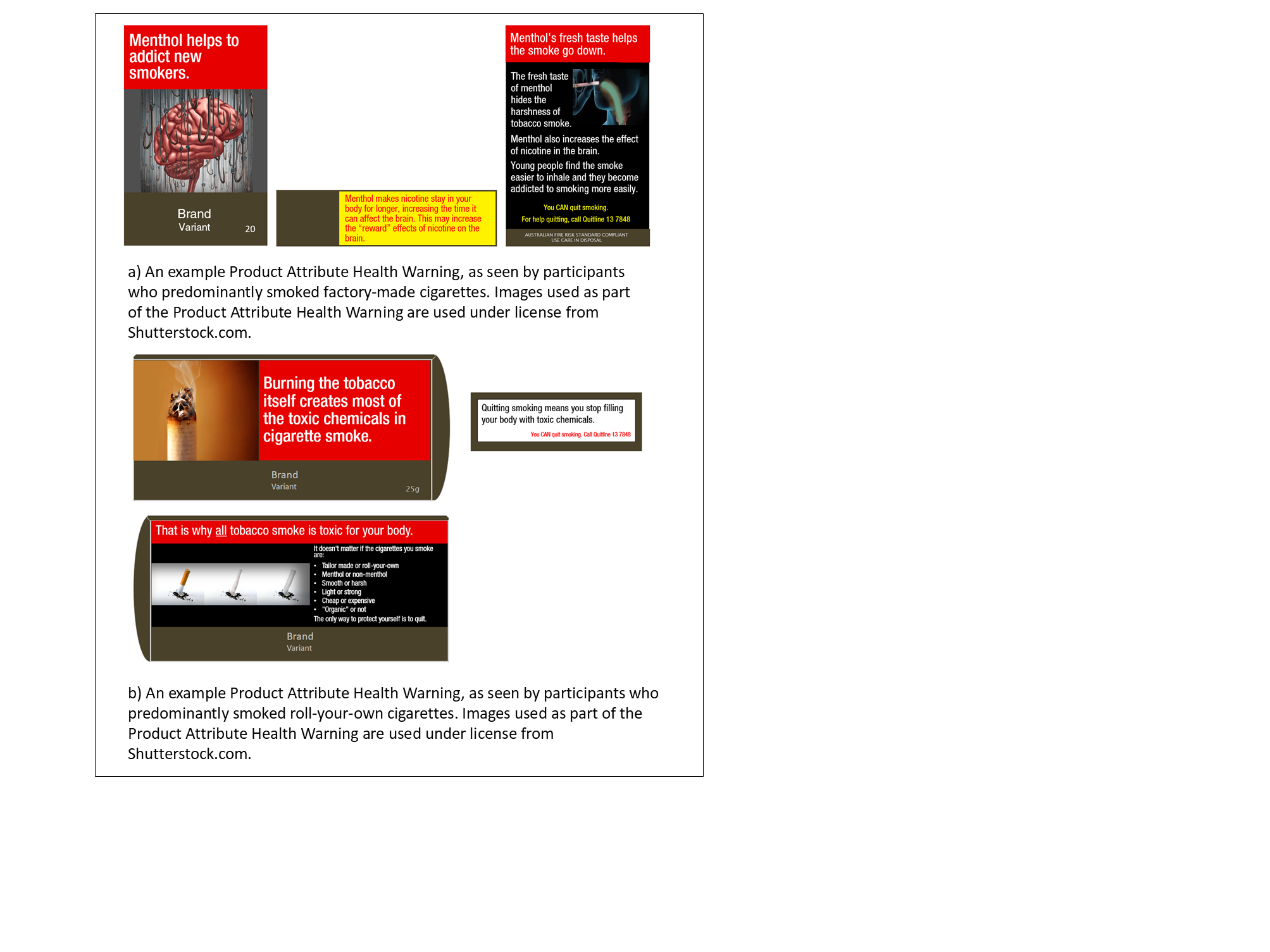
