## Supplemental Table 1 (S1) for "Do short-term responses to product manipulation health warnings predict subsequent quitting-related behaviours? Findings from an Australian cohort study"

**Table S1.** Socio-demographic and smoking characteristics among those who completed each survey.

|  | Completed baseline survey | Completed baseline survey and 8-day follow-up | Completed baseline survey and 8-day follow-up only | Completed baseline survey, 8-day follow-up, and 4-week follow-up | ꭕ^2^ test |
| --- | --- | --- | --- | --- | --- |
|  | n (%) | n (%) | n (%) | n (%) | *p*-value |
| *Total* | 1,295 (100.0) | 712 (100.0) | 411 (100.0) | 301 (100.0) |  |
| *Age group* |  |  |  |  | 0.151 |
| 18-39 years | 767 (59.2) | 396 (55.6) | 238 (57.9) | 158 (52.5) |  |
| 40-69 years | 528 (40.8) | 316 (44.4) | 173 (42.1) | 143 (47.5) |  |
| *Gender ^a^* |  |  |  |  | 0.514 |
| Man / male | 644 (49.7) | 309 (43.4) | 174 (42.3) | 135 (44.9) |  |
| Woman / female | 646 (49.9) | 400 (56.2) | 235 (57.2) | 165 (54.8) |  |
| Another term | 3 (0.2) | 1 (0.1) | 1 (0.2) | 0 (0.0) |  |
| Prefer not to say | 2 (0.2) | 2 (0.3) | 1 (0.2) | 1 (0.3) |  |
| *Highest level of education* |  |  |  |  | 0.598 |
| No tertiary education | 725 (56.0) | 428 (60.1) | 243 (59.1) | 185 (61.5) |  |
| Tertiary education | 562 (43.4) | 279 (39.2) | 164 (39.9) | 115 (38.2) |  |
| *Socio-economic area* |  |  |  |  | 0.477 |
| Low (1-40%) | 505 (39.0) | 293 (41.2) | 173 (42.1) | 120 (39.9) |  |
| Mid (41-80%) | 504 (38.9) | 287 (40.3) | 168 (40.9) | 119 (39.5) |  |
| High (81-100%) | 286 (22.1) | 132 (18.5) | 70 (17.0) | 62 (20.6) |  |
| *Geographic region* |  |  |  |  | 0.859 |
| Metropolitan | 927 (71.6) | 504 (70.8) | 292 (71.1) | 212 (70.4) |  |
| Regional | 368 (28.4) | 208 (29.2) | 119 (29.0) | 89 (29.6) |  |
| *Aboriginal and/or Torres Strait Islander ^b^* |  |  |  |  | 0.326 |
| No | 1,199 (92.6) | 673 (94.5) | 386 (93.9) | 287 (95.4) |  |
| Yes | 80 (6.2) | 35 (4.9) | 23 (5.6) | 12 (4.0) |  |
| Prefer not to say | 16 (1.2) | 4 (0.6) | 2 (0.5) | 2 (0.7) |  |
| *Health Care Card or Pensioner Concession Card holder* |  |  |  |  | 0.109 |
| No | 857 (66.2) | 449 (63.1) | 249 (60.6) | 200 (66.5) |  |
| Yes | 438 (33.8) | 263 (36.9) | 162 (39.4) | 101 (33.6) |  |
| *Predominant product use* |  |  |  |  | <0.001 |
| TM cigarettes | 717 (55.4) | 391 (54.9) | 252 (61.3) | 139 (46.2) |  |
| RYO cigarettes | 263 (20.3) | 160 (22.5) | 91 (22.1) | 69 (22.9) |  |
| Menthol cigarettes | 315 (24.3) | 161 (22.6) | 68 (16.6) | 93 (30.9) |  |
| *Number of cigarettes smoked per day* |  |  |  |  | 0.756 |
| <10 | 661 (51.0) | 344 (48.3) | 197 (47.9) | 147 (48.8) |  |
| 10+ | 585 (45.2) | 345 (48.5) | 199 (48.4) | 146 (48.5) |  |
| *Quit attempts in past year* |  |  |  |  | 0.889 |
| None | 604 (46.6) | 318 (44.7) | 182 (44.3) | 136 (45.2) |  |
| At least once | 610 (47.1) | 354 (49.7) | 207 (50.4) | 147 (48.8) |  |
| Don’t know/can’t say | 81 (6.3) | 40 (5.6) | 22 (5.4) | 18 (6.0) |  |
| *Intention to quit in next 30 days* |  |  |  |  | 0.939 |
| No | 1,020 (78.8) | 562 (78.9) | 324 (78.8) | 238 (79.1) |  |
| Yes | 275 (21.2) | 150 (21.1) | 87 (21.2) | 63 (20.9) |  |
| *Frequency of e-cigarette use* |  |  |  |  | 0.384 |
| Less than monthly | 742 (57.3) | 441 (61.9) | 249 (60.6) | 192 (63.8) |  |
| At least monthly | 553 (42.7) | 271 (38.1) | 162 (39.4) | 109 (36.2) |  |
| *Condition* |  |  |  |  | 0.255 |
| PAHW | 651 (50.3) | 356 (50.0) | 198 (48.2) | 158 (52.5) |  |
| PAHW+Video | 644 (49.7) | 356 (50.0) | 213 (51.8) | 143 (47.5) |  |

Notes: Proportions are rounded so may not sum to 100%. Among those who completed the baseline survey, no response was provided for highest level of education for n=8 participants and no response was provided for number of cigarettes smoked per day for n=49 participants. Chi-square tests were conducted to assess significant differences in socio-demographic and smoking characteristics between the sample of participants who completed the baseline survey and 8-day follow-up only (column 4) and participants who completed the baseline survey, 8-day follow-up and 4-week follow-up (column 5).

Abbreviations: TM=tailor-made; RYO=roll-your-own; PAHW=product attribute health warning.

^a^ Chi-square test was conducted with ‘another term’ and ‘prefer not to say’ coded as missing due to limited observations.

^b^ Chi-square test was conducted with ‘prefer not to say’ coded as ‘no’ due to limited observations.
