## Supplemental Table 2 (S2) for "Do short-term responses to product manipulation health warnings predict subsequent quitting-related behaviours? Findings from an Australian cohort study"

Table S2. Results from binary logistic regression models assessing predictive validity of PAHW response measures for quitting-related behavioural outcomes, including 8-day measures of the quitting-related outcomes as covariates.

|  | All three smoke-limiting behaviours  (N=282) | | At least one quit attempt  (N=292) | |
| --- | --- | --- | --- | --- |
|  | Adj. Odds Ratio (95% CI) | *p*-value | Adj. Odds Ratio (95% CI) | *p*-value |
| Knowledge of industry manipulation of cigarettes | 1.24 (0.84, 1.83) | 0.287 | 1.43 (0.93, 2.18) | 0.100 |
| Industry-centric negative emotional responses | 1.61 (1.16, 2.23) | 0.004 | 1.23 (0.89, 1.70) | 0.211 |
| Product-specific smoking dissonance | 1.81 (1.21, 2.71) | 0.004 | 2.14 (1.40, 3.26) | <0.001 |

Note: All models adjusted for intentions to quit in the next 30 days measured at baseline, number of days between 8-day and 4-week follow-up surveys, age group, number of cigarettes smoked per day, predominant product use, and frequency of e-cigarette use. Models pertaining to smoke-limiting behaviours also adjusted for region, and models pertaining to quit attempts also adjusted for gender. Analysis was not conducted for models pertaining to 7-day sustained abstinence as this measure was not collected at the 8-day follow-up survey.
